# Monovalent XBB.1.5 COVID-19 vaccine effectiveness against hospitalisations and deaths during the Omicron BA.2.86/JN.1 period among older adults in seven European countries: A VEBIS-EHR Network Study

**DOI:** 10.1101/2024.07.04.24309832

**Authors:** Baltazar Nunes, James Humphreys, Nathalie Nicolay, Toon Braeye, Izaak Van Evercooren, Christian Holm Hansen, Ida Rask Moustsen-Helms, Chiara Sacco, Massimo Fabiani, Jesús Castilla, Iván Martínez-Baz, Hinta Meijerink, Ausenda Machado, Patricia Soares, Rickard Ljung, Nicklas Pihlström, Anthony Nardone, Sabrina Bacci, Susana Monge

**Affiliations:** Epiconcept, Paris, France; Vaccine Preventable Diseases and Immunisation, European Centre for Disease Prevention and Control (ECDC), Solna, Sweden; Sciensano, Juliette Wytsmanstraat 14. 1050 Elsene, Belgium; Department of Infectious Disease Epidemiology and Prevention, Statens Serum Institut, Copenhagen, Denmark; Infectious Diseases Department, Istituto Superiore di Sanità, Rome, Italy; European Programme on Intervention Epidemiology Training (EPIET), European Centre for Disease Prevention and Control, Stockholm, Sweden; Instituto de Salud Pública de Navarra – IdiSNA, Pamplona, Spain; CIBER on Epidemiology and Public Health, Spain; Norwegian Institute of Public Health (NIPH), Oslo, Norway; Instituto Nacional de Saúde Doutor Ricardo Jorge, Lisboa, 1600-609, Portugal; Swedish Medical Products Agency, Uppsala, Sweden; Department of Communicable Diseases, National Centre of Epidemiology, Institute of Health Carlos III, Madrid, Spain; CIBER on Infectious Diseases, Spain

**Keywords:** COVID-19, SARS-CoV-2, vaccine effectiveness, hospitalisation, cohort design, electronic health records, multi-country study

## Abstract

**Background:** Monovalent XBB.1.5 vaccine was administered among those aged ≥65 years in EU/EEA countries in autumn 2023; soon after SARS-Cov-2 BA.2.86/JN.1 lineages became dominant. We aimed to estimate XBB.1.5 vaccine effectiveness (VE) against COVID-19-related hospitalisations and deaths during a period of BA.2.86/JN.1 predominance using a European multi-country study.

**Methods:** We linked electronic health record data to create historical cohorts in Belgium, Denmark, Italy, Navarre (Spain), Norway, Portugal and Sweden. We included individuals aged ≥65 years eligible for the autumnal 2023 COVID-19 vaccine with at least a primary series. Follow-up started when ≥80% of country-specific sequenced viruses were BA.2.86/JN.1 lineages (4/12/23 to 08/01/24) and ended 25/02/2024. At study site level, we estimated the overall vaccine confounder-adjusted (for age, sex, country’s region, comorbidities and previous booster doses) hazard ratio (aHR) of COVID-19 hospitalisations and deaths between individuals with ≥14 days after vaccination and individuals unvaccinated in autumn 2023, as well as by time since vaccination and stratified by age groups. VE was estimated as (1-pooled aHR)x100 with a random effects model.

**Results:** XBB.1.5 VE against COVID-19 hospitalisations was 50% (95%CI: 45 to 55) and 41% (95%CI: 35 to 46) in 65-79-year-olds and in ≥80-year-olds respectively. VE against COVID19-related-death was 58% (95%CI: 42 to 69) and 48% (95%CI: 38 to 57), respectively, in both age groups. VE estimates against each respective outcome declined in all age group over time.

**Conclusion:** Monovalent XBB.1.5 vaccine had a moderate protective effect against severe COVID-19 likely caused by BA.2.86/JN.1 during the 2023/2024 winter, among persons aged ≥65.

## Introduction

In autumn 2023, COVID-19 vaccination campaigns were carried out in EU/EEA countries aiming to reduce the risk of severe disease among the most vulnerable populations. The target groups generally included those aged ≥60 or ≥65 years, persons living with comorbidities or conditions that could increase the risk of severe disease, health professionals, and caregivers. The monovalent Omicron XBB.1.5 vaccine was used in most EU/EEA countries and delivered as a booster dose or primary vaccination during autumn/winter 2023/24. In January 2024 (1) the median vaccine coverage among EU/EEA countries was 12% (range 0.01%–66.1%) and 17.1 (range 0.01%–89.3%), for those aged ≥60 or ≥80 years respectively - a range which indicates variability between countries in terms of vaccine uptake. Eighty-two percent of the vaccines administered during the autumn/winter 2023/24 campaign were Comirnaty Omicron XBB.1.5 (Pfizer BioNTech) vaccine (1).

In early autumn 2023, in EU/EEA countries, the XBB.1.5 lineage was predominant. The BA.2.86 and JN.1 SARS-CoV-2 lineages rapidly increased, gaining dominance in all EU/EEA countries by mid-December (2).

Several studies reported XBB.1.5 vaccine effectiveness (VE) against severe disease in the first months of the autumn/winter season at a time of XBB.1.5 predominance (3–6), including one study conducted within the Vaccine Effectiveness Burden and Impact Studies project EHR network (VEBIS-EHR) (4). These studies reported VE against COVID-19 hospitalisations ranging between 62% to 74%, 52% to 74%, and 52% to 66% among those aged ≥18, ≥65 and ≥80 years old respectively.

The BA.2.86 SARS-CoV-2 lineage presents more than 30 amino acid mutations compared to the XBB.1.5 lineage, and JN.1, a descendant of BA.2.86, harbours the L455S substitution at the receptor binding site. These mutations could provide immune escape capacities and reduce the effectiveness of the XBB.1.5 vaccine (7). However, studies of XBB.1.5 VE against severe disease during periods of BA.2.86 or JN.1 predominance are still scarce (5,8). In this context, we aimed to estimate XBB.1.5 VE against COVID-19-related hospitalisations and deaths overall and by time since vaccination during a period of BA.2.86/JN.1 predominance within a European multi-country study among older adults aged 65-79 and ≥80 years old.

## Methods

This study was developed under the VEBIS-EHR network. The VEBIS project, funded by the European Centre for Prevention and Disease Control (ECDC), aims to monitor COVID-19 VE using linkage of electronic health records (EHR) within an EU/EEA multi-country setting. As of April 2024, seven countries participate in the VEBIS-EHR network: Belgium, Denmark, Italy, Spain (Navarre), Norway, Portugal, and Sweden.

Detailed methods and the results of VEBIS-EHR studies have been published previously (9–11). Briefly, using a common protocol (12) we developed a historical cohort study by reconstructing study site cohorts of individuals aged 65 years or older resident in the and eligible to receive the autumnal 2023 vaccine dose at the start of the country-specific vaccination campaign (Supplementary material, Table S1, Annex 1), i.e., those aged 65 years or older, with primary vaccination series completed at least 180 days ago and who, in the last 90 days, had no vaccine dose administered nor documented SARS-CoV-2 infection or COVID-19 hospitalisation. Detailed information on the eligibility criteria is presented in the Supplementary material, Annex 2. Vaccination status, baseline characteristics, and outcome occurrence were obtained by linking national/regional EHR. We defined BA.2.86/JN.1 lineage predominance period at the study-site level, designating the period of predominance to have started by the date on which 80% of sequenced samples were attributable to BA.2.86/JN.1 based on data extracted from the ECDC European Respiratory Virus Surveillance Summary (ERVISS)(13) database and subset to the countries of interest.

If the number of the weekly sequenced viruses was low, BA.2.86 frequency data from neighbouring countries were used as a proxy (Supplementary material, Annex 1). In Table S1 we present details on the start of BA.2.86 predominance period by study site. The end of the study period was 25 February in all study sites, approximately two months before data extraction (April 2024) to ensure EHR were sufficiently updated.

Among those identified as eligible at the start of the vaccination campaign (Supplementary material, Annex 2), we excluded all individuals hospitalised due to COVID-19 between the start of the vaccination campaign and the country-specific starting date of the study. Individual follow-up started at the beginning of the country-specific study period and ceased at the date of either the outcome, death for any cause, or the end of the study period - whichever was earliest. Vaccination status was treated as a time-dependent exposure, excluding the time-interval 1-14 days post-vaccination from the analysis.

Hospitalisation due to COVID-19 was defined as a hospital admission due to a severe acute respiratory infection with a SARS-CoV-2 positive test from 14 days before to one day after admission, or with COVID-19 as the primary diagnosis in admission on discharge records. A COVID-19-related death was defined as a death with COVID-19 coded as cause, or death for any cause with a SARS-CoV-2 positive test in the 30 days preceding death.

We undertook a two-stage pooling of estimates calculated at the study-site level. In the first stage, site-specific estimates of confounder-adjusted vaccine hazard ratios (aHR) and 95% confidence intervals (95%CI) were calculated using Cox regression with calendar time as the underlying scale adjusted by 5-year age groups, sex, region in the country, comorbidities, number of vaccine booster doses received prior to the current vaccination campaign, and nationality and socioeconomic status in some of the study sites (Supplementary material, Annex 3 and 4). In the second stage, study site-specific aHR estimates and standard errors were pooled using a random-effects meta-analysis using Paule-Mantel method. Pooled VE was estimated as (1-pooled aHR)x100. To assess heterogeneity, we used the I^2 index (14). A fixed-effects model was used as a secondary analysis.

## Results

At the end of the individual observation period, we included 13,907,924 and 6,533,724 persons across both age groups, in the unvaccinated and vaccinated cohorts, respectively. Among the vaccinated cohorts, at the end of the observation period, 2,604,152 (39.9%) and 3,929,572 (60.1%) belonged, respectively, to those who were vaccinated 14 to 89 days and 90 to 179 days.

The proportion of individuals with high-risk comorbidities (7.3% vs 2.6%) and with a greater number of previous booster doses (≥2 boosters: 92.3% vs 33.1%) was higher among vaccinated than unvaccinated. Further, but to a lesser extent, vaccinated persons with a longer time since vaccination (90-179 days) also had a greater frequency of high-risk comorbidities (9.0% vs 4.8%) and a greater number of previous boosters (≥2 boosters: 94.3% vs 89.4%) doses compared to those vaccinated with less time since vaccination (14-90 days) (Table 1).

**Table 1.**
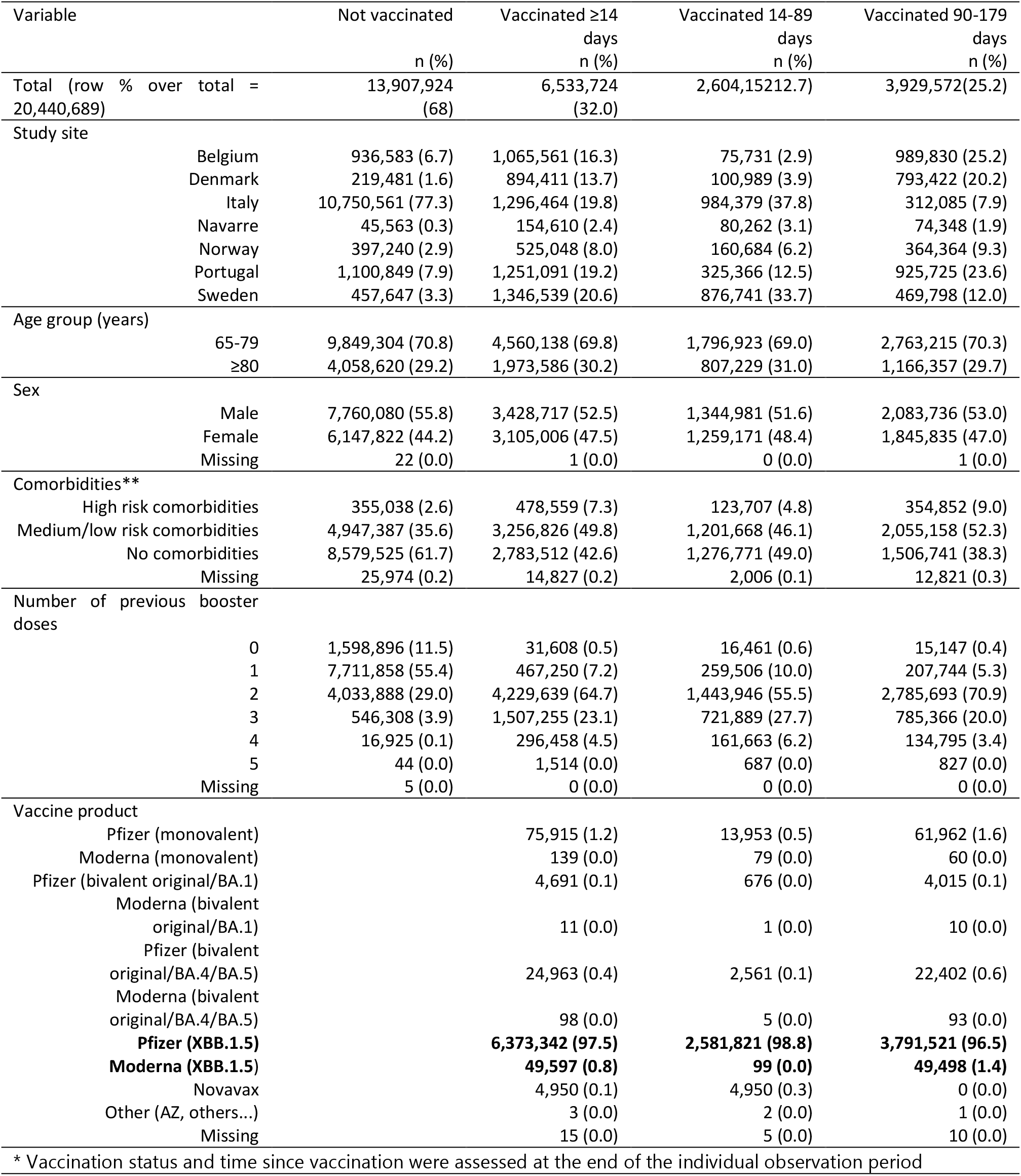
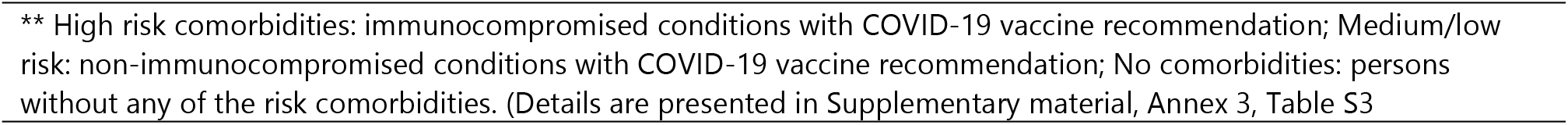
Descriptive characteristics of the study population (N= 20,441,648) by vaccination status and time since vaccination at the end of the study period*, within the seven study sites (Belgium, Denmark, Italy, Navarre-Spain, Norway, Portugal, and Sweden during BA.2.86/JN.1 predominant period (from 4 December 2023 until 25 February 2024): VEBIS-EHR network.

| Variable |  | Not vaccinated | Vaccinated ≥14 days | Vaccinated 14-89 days | Vaccinated 90-179 days |
| --- | --- | --- | --- | --- | --- |
|  |  | n (%) | n (%) | n (%) | n (%) |
| Total (row % over total = 20,440,689) |  | 13,907,924 (68) | 6,533,724 (32.0) | 2,604,152 (12.7) | 3,929,572 (25.2) |
| Study site |  |  |  |  |  |
|  | Belgium | 936,583 (6.7) | 1,065,561 (16.3) | 75,731 (2.9) | 989,830 (25.2) |
|  | Denmark | 219,481 (1.6) | 894,411 (13.7) | 100,989 (3.9) | 793,422 (20.2) |
|  | Italy | 10,750,561 (77.3) | 1,296,464 (19.8) | 984,379 (37.8) | 312,085 (7.9) |
|  | Navarre | 45,563 (0.3) | 154,610 (2.4) | 80,262 (3.1) | 74,348 (1.9) |
|  | Norway | 397,240 (2.9) | 525,048 (8.0) | 160,684 (6.2) | 364,364 (9.3) |
|  | Portugal | 1,100,849 (7.9) | 1,251,091 (19.2) | 325,366 (12.5) | 925,725 (23.6) |
|  | Sweden | 457,647 (3.3) | 1,346,539 (20.6) | 876,741 (33.7) | 469,798 (12.0) |
| Age group (years) |  |  |  |  |  |
|  | 65-79 | 9,849,304 (70.8) | 4,560,138 (69.8) | 1,796,923 (69.0) | 2,763,215 (70.3) |
|  | ≥80 | 4,058,620 (29.2) | 1,973,586 (30.2) | 807,229 (31.0) | 1,166,357 (29.7) |
| Sex |  |  |  |  |  |
|  | Male | 7,760,080 (55.8) | 3,428,717 (52.5) | 1,344,981 (51.6) | 2,083,736 (53.0) |
|  | Female | 6,147,822 (44.2) | 3,105,006 (47.5) | 1,259,171 (48.4) | 1,845,835 (47.0) |
|  | Missing | 22 (0.0) | 1 (0.0) | 0 (0.0) | 1 (0.0) |
| Comorbidities** |  |  |  |  |  |
|  | High risk comorbidities | 355,038 (2.6) | 478,559 (7.3) | 123,707 (4.8) | 354,852 (9.0) |
|  | Medium/low risk comorbidities | 4,947,387 (35.6) | 3,256,826 (49.8) | 1,201,668 (46.1) | 2,055,158 (52.3) |
|  | No comorbidities | 8,579,525 (61.7) | 2,783,512 (42.6) | 1,276,771 (49.0) | 1,506,741 (38.3) |
|  | Missing | 25,974 (0.2) | 14,827 (0.2) | 2,006 (0.1) | 12,821 (0.3) |
| Number of previous booster doses |  |  |  |  |  |
|  | 0 | 1,598,896 (11.5) | 31,608 (0.5) | 16,461 (0.6) | 15,147 (0.4) |
|  | 1 | 7,711,858 (55.4) | 467,250 (7.2) | 259,506 (10.0) | 207,744 (5.3) |
|  | 2 | 4,033,888 (29.0) | 4,229,639 (64.7) | 1,443,946 (55.5) | 2,785,693 (70.9) |
|  | 3 | 546,308 (3.9) | 1,507,255 (23.1) | 721,889 (27.7) | 785,366 (20.0) |
|  | 4 | 16,925 (0.1) | 296,458 (4.5) | 161,663 (6.2) | 134,795 (3.4) |
|  | 5 | 44 (0.0) | 1,514 (0.0) | 687 (0.0) | 827 (0.0) |
|  | Missing | 5 (0.0) | 0 (0.0) | 0 (0.0) | 0 (0.0) |
| Vaccine product |  |  |  |  |  |
|  | Pfizer (monovalent) |  | 75,915 (1.2) | 13,953 (0.5) | 61,962 (1.6) |
|  | Moderna (monovalent) |  | 139 (0.0) | 79 (0.0) | 60 (0.0) |
|  | Pfizer (bivalent original/BA.1) |  | 4,691 (0.1) | 676 (0.0) | 4,015 (0.1) |
|  | Moderna (bivalent original/BA.1) |  | 11 (0.0) | 1 (0.0) | 10 (0.0) |
|  | Pfizer (bivalent original/BA.4/BA.5) |  | 24,963 (0.4) | 2,561 (0.1) | 22,402 (0.6) |
|  | Moderna (bivalent original/BA.4/BA.5) |  | 98 (0.0) | 5 (0.0) | 93 (0.0) |
|  | **Pfizer (XBB.1.5)** |  | **6,373,342 (97.5)** | **2,581,821 (98.8)** | **3,791,521 (96.5)** |
|  | **Moderna (XBB.1.5)** |  | **49,597 (0.8)** | **99 (0.0)** | **49,498 (1.4)** |
|  | Novavax |  | 4,950 (0.1) | 4,950 (0.3) | 0 (0.0) |
|  | Other (AZ, others...) |  | 3 (0.0) | 2 (0.0) | 1 (0.0) |
|  | Missing |  | 15 (0.0) | 5 (0.0) | 10 (0.0) |

\* Vaccination status and time since vaccination were assessed at the end of the individual observation period
\*\* High risk comorbidities: immunocompromised conditions with COVID-19 vaccine recommendation; Medium/low risk: non-immunocompromised conditions with COVID-19 vaccine recommendation; No comorbidities: persons without any of the risk comorbidities. (Details are presented in Supplementary material, Annex 3, Table S3)

In the 65-79 years age group there were 1,765 hospitalisation events among 17.8 million person-months at risk in unvaccinated, and 908 COVID-19 hospitalisations events among 9.9 million person-months at risk in vaccinated persons. XBB.1.5 COVID-19 VE against COVID-19 hospitalisations was 50% (95%CI: 45 to 55) overall, 51% (95%CI: 45 to 56) and 47% (95%CI: 32 to 59) among those who received the vaccine respectively between 14 to 89 days and 90 to 179 days.

In ≥80 years age-group, there were 2,130 COVID-19 hospitalisation events among 7.0 million person-months at risk in unvaccinated, and 1,306 among 4.1 million person-months at risk in vaccinated persons. VE was 41% (95%CI: 35 to 46) overall and 42% (95%CI: 36 to 47) and 38% (95%CI: 17 to 54%) by time since vaccination among those who received the vaccine between 14 to 89 days and 90 to 179 days, respectively.

In the 65–79-year age-group there were 256 COVID-19 related deaths among 15.9 million person-months at risk in unvaccinated, and 151 among 7.8 million person-months at risk in vaccinated individuals. XBB.1.5 COVID-19 VE estimates against COVID-19-related deaths, was 58% (95%CI: 42 to 69) overall, 59% (95%CI: 41 to 72) and 54% (95%CI: −17 to 82) respectively among those who received the vaccine between 14 to 89 days and 90 to 179 days.

In the ≥80-year age-group, there were 570 COVID-19 related deaths event among 6.6 million person-months at risk in unvaccinated, and 389 among 3.4 million person-months at risk in unvaccinated persons. The XBB.1.5 COVID-19 was 48% (95%CI: 38 to 57) overall, 51% (95%CI: 42 to 59) and 9% (95%CI: −86 to 56%) among those who received the vaccine between 14 to 89 days and 90 to 179 days, respectively (Table 2).

**Table 2.**
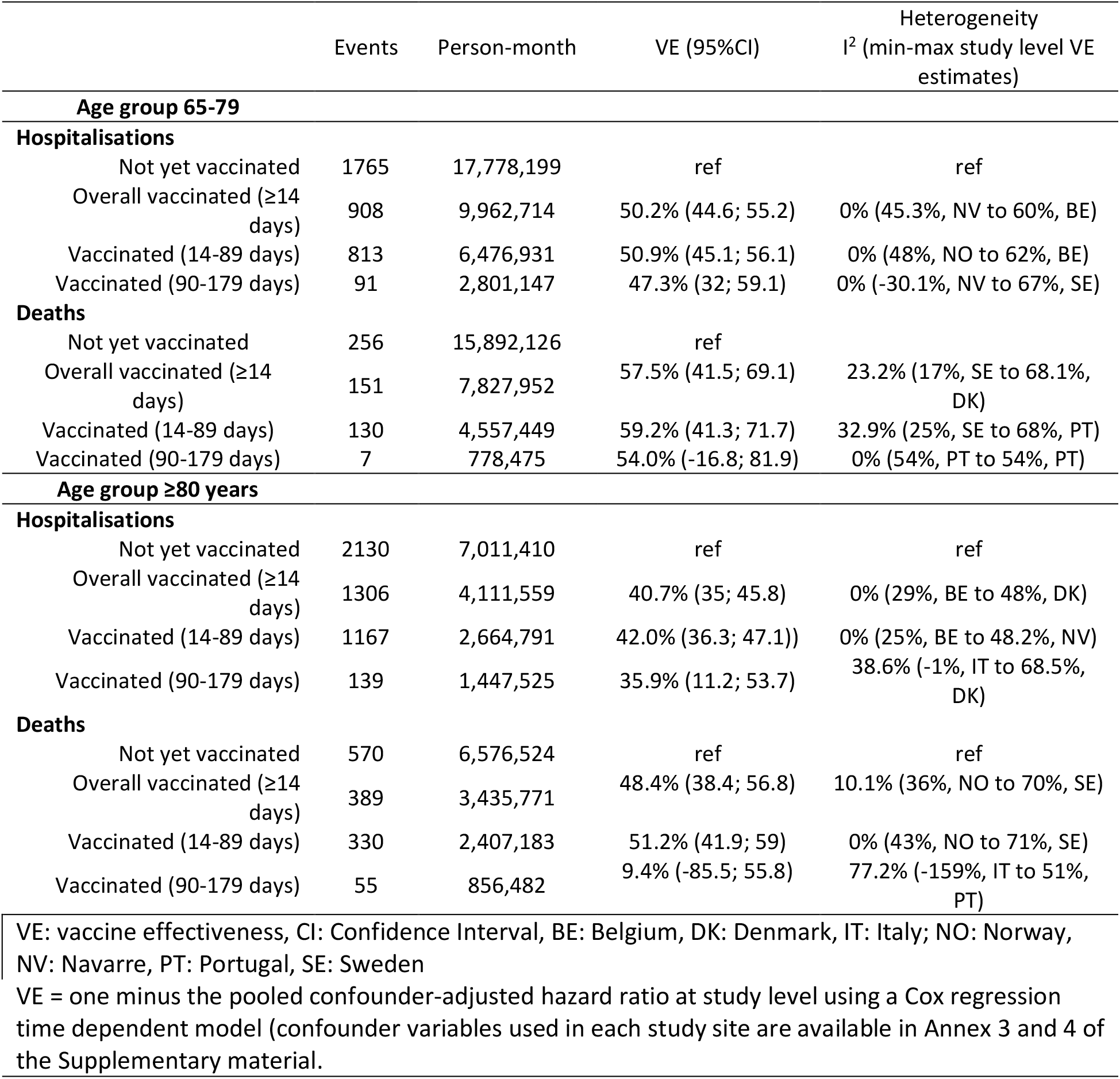
Number of events (COVID-19 hospitalisations or COVID-19 related deaths), person months at risk by vaccine status, and vaccine effectiveness overall and by time since vaccination for individuals aged 65-79 and ≥80 years old, within the seven study sites (Belgium, Denmark, Italy, Navarre-Spain, Norway, Portugal, and Sweden), during BA.2.86/JN.1 predominance period up to 25 February 2024, VEBIS-EHR network

|  | Events | Person-month | VE (95%CI) | Heterogeneity<br>I<sup>2</sup> (min-max study level VE estimates) |
| --- | --- | --- | --- | --- |
| **Age group 65-79** |  |  |  |  |
| **Hospitalisations** |  |  |  |  |
| Not yet vaccinated | 1765 | 17,778,199 | ref | ref |
| Overall vaccinated (≥14 days) | 908 | 9,962,714 | 50.2% (44.6; 55.2) | 0% (45.3%, NV to 60%, BE) |
| Vaccinated (14-89 days) | 813 | 6,476,931 | 50.9% (45.1; 56.1) | 0% (48%, NO to 62%, BE) |
| Vaccinated (90-179 days) | 91 | 2,801,147 | 47.3% (32; 59.1) | 0% (-30.1%, NV to 67%, SE) |
| **Deaths** |  |  |  |  |
| Not yet vaccinated | 256 | 15,892,126 | ref |  |
| Overall vaccinated (≥14 days) | 151 | 7,827,952 | 57.5% (41.5; 69.1) | 23.2% (17%, SE to 68.1%, DK) |
| Vaccinated (14-89 days) | 130 | 4,557,449 | 59.2% (41.3; 71.7) | 32.9% (25%, SE to 68%, PT) |
| Vaccinated (90-179 days) | 7 | 778,475 | 54.0% (-16.8; 81.9) | 0% (54%, PT to 54%, PT) |
| **Age group ≥80 years** |  |  |  |  |
| **Hospitalisations** |  |  |  |  |
| Not yet vaccinated | 2130 | 7,011,410 | ref | ref |
| Overall vaccinated (≥14 days) | 1306 | 4,111,559 | 40.7% (35; 45.8) | 0% (29%, BE to 48%, DK) |
| Vaccinated (14-89 days) | 1167 | 2,664,791 | 42.0% (36.3; 47.1)) | 0% (25%, BE to 48.2%, NV) |
| Vaccinated (90-179 days) | 139 | 1,447,525 | 35.9% (11.2; 53.7) | 38.6% (-1%, IT to 68.5%, DK) |
| **Deaths** |  |  |  |  |
| Not yet vaccinated | 570 | 6,576,524 | ref | ref |
| Overall vaccinated (≥14 days) | 389 | 3,435,771 | 48.4% (38.4; 56.8) | 10.1% (36%, NO to 70%, SE) |
| Vaccinated (14-89 days) | 330 | 2,407,183 | 51.2% (41.9; 59) | 0% (43%, NO to 71%, SE) |
| Vaccinated (90-179 days) | 55 | 856,482 | 9.4% (-85.5; 55.8) | 77.2% (-159%, IT to 51%, PT) |
| VE: vaccine effectiveness, CI: Confidence Interval, BE: Belgium, DK: Denmark, IT: Italy; NO: Norway, NV: Navarre, PT: Portugal, SE: Sweden |  |  |  |  |
| VE = one minus the pooled confounder-adjusted hazard ratio at study level using a Cox regression time dependent model (confounder variables used in each study site are available in Annex 3 and 4 of the Supplementary material). |  |  |  |  |

There was low to moderate heterogeneity between study sites’ aHR estimates among pooled VE estimates (I^2^=0% to 77%). High heterogeneity (I^2^=77%) was observed in the VE estimate against death within 90-179 days since vaccination for the 80+ cohort, leading to a very wide 95% confidence interval for this estimate. Fixed effects pooled estimates were similar to the random effect estimates presented in the primary analysis (Supplementary figures – Annex 6).

## Discussion

Our results indicate that in the population aged 65 years and older, the 2023 autumnal monovalent XBB.1.5 booster conferred protection against severe COVID-19 outcomes during the period in which COVID-19 cases were likely to be caused by the Omicron BA.2.86 or JN.1 lineages. These estimates of protection represent a reduction in risk that ranged from 42% to 51% for COVID-19 hospitalisation and from 51 to 59% for COVID-19 deaths for those who received the vaccine in the last three months, and were consistently lower among those aged ≥80 years. For those vaccinated, but with more time since vaccination (three or more months) during the BA.2.86/JN.1 predominance period, XBB.1.5 vaccine presented lower levels of protection against COVID-19 hospitalisations, ranging from 38% to 47%, and against COVID-19 related deaths ranging from 9% to 54% though, as discussed earlier, the latter estimates against COVID-19-related deaths had low precision.

Our results suggest a lower VE among those who received a vaccine between 14 and 89 days in the BA.2.86/JN.1 predominant period, compared to the XBB predominant period estimates obtained in the VEBIS-EHR network (4). Specifically, in the 65–79-year-old XBB.1.5 VE against hospitalisations decreased from 64% (95%CI 55 to 72) in the XXB period to 51% (95%CI 45 to 56) in the BA 2.86/JN1 period and for those ≥80 years from 65% (95%CI 56 to 71) to 42% (95%CI 36 to 47). The XBB.1.5 VE against COVID-19-related deaths declined in those aged 65-79 years from 67% (95%CI: 42 to 81) to 59% (95%CI 41 to 72) and in those ≥80 years from 67% (95%CI: 41 to 81) to 51% (95%CI: 42 to 59). These differences could be due to higher natural immunity in the comparison group due to recent SARS-CoV-2 infection, although other studies employing different study designs have also found evidence of reduced vaccine effectiveness against BA.2.86/JN.1 relative to XBB.1.5 (15).

Our results are concordant with results from other studies that estimated COVID-19 VE against severe outcomes potentially due to XBB sub-lineages or JN.1 SARS-CoV-2 lineages. A test-negative design study conducted in England (8) in the population aged 65 years or more showed a decrease in VE against hospitalisations from 74% (95%CI: 62% to 82%) against XBB sub-lineages to 37% (95%CI: −20 to 66) against JN.1, for those who were vaccinated 2-4 weeks after vaccination before. Similar results were also observed in two studies conducted in the United States (5,6) that aimed to estimate XBB.1.5 VE against hospitalisations likely due to XBB sub-lineages versus JN.1 among individuals aged ≥18 years. In both studies, a reduction in the protection of XBB.1.5 vaccine against COVID-19 hospitalisations was observed within the first 60 days after vaccination, respectively from 62% (95%CI: 44% to 74%) to 32% (95%CI: 3% to 52%) (5), and from 74% (95%CI: 49% to 87%) to 50% (95%CI: 15% to 71%) (6).

The results presented here should be interpreted cautiously, considering the potential presence of confounding bias, given its observational nature, and the possible misclassification of the vaccine status and outcome once we used an EHR-based multicentre study. The vaccine status hazard ratios were adjusted for several potential confounding factors at the study site level. Still, although this adjustment was made, we cannot exclude the presence of confounding residual bias from our analysis. Considering misclassification of the exposure, vaccination information was extracted from national vaccination registries and measured prior to, and independently of, the outcome. Estimates of 2023 autumnal COVID-19 vaccine coverage observed within our EHR-based study population were very close to the officially reported equivalents produced by the ECDC (Supplementary, Annex 6, Table S5). We considered our VE estimate representative of XBB.1.5 vaccine performance given that 98.3% (Table 1) of the vaccines received during the study period were the monovalent XBB.1.5. Among these, 99% were the Pfizer Comirnaty XBB.1.5-adapted vaccine, which indicates that our VE estimates represent the effectiveness of this vaccine brand in general.

Additionally, we cannot rule out the presence of misclassification or underreporting of COVID-19 hospitalisations and deaths, even using national hospitalisations, death and laboratory registries given the lag between event and full recording of such events in the relevant EHRs, and also due the reduction in SARS-CoV-2 testing that would underestimate COVID-19 related deaths identification. To reduce the effect of misclassification bias due to unextracted events, or otherwise due to extraction of incomplete records, a two-month delay between the end of the study period (i.e., the last possible event) and extraction of data from source EHRs has been implemented. For the purpose of this analysis, no EHR extractions were undertaken until at least April 2024. Nevertheless, we do not expect that misclassification of the outcome could be differential by the vaccine status. If underreporting of events is present in our data, we do not expect a bias to arise from it, only a loss in the number of events and statistical power. On the other hand, if non-COVID-19 hospitalisation are classified as COVID-19 event, we expect that our estimates would be biased to the null effect (16).

Another issue to be addressed, will be the high diffusion of self-diagnosis through at-home testing and the possible under-ascertainment of asymptomatic and mild cases, it is likely that some SARS-CoV-2 infections were not reported, thus leading to a possible overestimation of the eligible individuals at the start of the vaccination campaigns. This was likely more frequent in those who did not receive the seasonal booster, possibly causing an underestimation of VE.

The comparison of VE by time since XBB.1.5 administration among different epidemic phases, like XXB vs BA.2.86 predominance periods, should be interpreted with cautions because of the possible different unmeasured characteristics among persons who received the seasonal booster at different times. For example, those who received the booster dose later might be generally less prone to adopt preventive measures and therefore have had a higher exposure to risky behaviours than those who received it earlier. Additionally, individuals with a recent infection will be higher among unvaccinated persons during BA.2.86/JN.1 period. Both these situations could have resulted in a relative underestimation of VE during the BA.2.86/JN.1 predominance period, and a slight overestimation of VE in the XBB predominance period.

Despite the limitations described above, our study also has several strengths. As we employed mostly national registries, large sample sizes and a large number of events were reported by study sites, leading to a high precision around VE estimates and allowing stratified VE estimates. Additionally, the fact that the study was conducted in seven different EU/EEA countries using a common scientific protocol published ahead of the study development (12), and the low to moderate heterogeneity observed between study-level VE estimates, reinforce the consistency and reproducibility of the results in the participating study sites.

In conclusion, our XBB.1.5 VE estimates against severe COVID-19, likely caused by BA.2.86/JN.1, were lower than those observed during the XBB.1.5 lineage predominance period. Nevertheless, the results of our study indicate that the XBB.1.5 vaccine offered moderate protection (≥40%) for up to six months after vaccination against severe COVID-19, likely caused by BA.2.86/JN.1, during the 2023/2024 winter in older (≥65 years old) populations.

## Acknowledgments

The authors would like to thank all the people from the study sites involved in the data collection and for producing estimates of VE, as without their work these results wouldn’t be available for the scientific community and the public.

We gratefully acknowledge all data contributors, i.e., the Authors and their Originating laboratories responsible for obtaining the specimens, and their Submitting laboratories for generating the genetic sequence and metadata and sharing via the GISAID Initiative, on which this research is based.

## Ethics

All study sites participating in this study conformed with their respective national and EU ethical and data protection requirements. Ethical statements for each of the participating study sites:

### Belgium

Data linkage and collection within the data-warehouse have been approved by the information security committee. The study was conducted in accordance with the Declaration of Helsinki. Ethical approval was granted for the gathering of data from hospitalized patients by the Committee for Medical Ethics from the Ghent University Hospital (reference number BC-07507) and authorization for possible individual data linkage using the national register number from the Information Security Committee (ISC) Social Security and Health (reference number IVC/KSZG/20/384). Linkage of hospitalized patient data to vaccination and testing within the LINK-VACC project was approved by the Medical Ethics Committee UZ Brussels–VUB on 3 February 2021 (reference number 2020/523), and authorization from the ISC Social Security and Health (reference number IVC/KSZG/21/034).

### Denmark

Only administrative register data was used for the study. According to Danish law, ethics approval is exempt for such research, and the Danish Data Protection Agency, which is dedicated ethics and legal oversight body, thus waives ethical approval for the study of administrative register data when no individual contact of participants is necessary, and only aggregate results are included as findings. The study is, therefore, fully compliant with all legal and ethical requirements, and there are no further processes available regarding such studies.

### Navarre (Spain)

The study was approved by Navarre’s Ethical Committee for Clinical Research, which waived the requirement of obtaining informed consent.

### Norway

Ethical approval was granted by Regional Committees for Medical and Health Research Ethics (REC) Southeast (reference number 122745). The Norwegian Institute of Public Health has performed a Data Protection Impact Assessment (DPIA) for Beredt C19.

### Portugal

The study received approval from the Ethical Committee and the Data Protection Officer of the Instituto Nacional de Saúde Doutor Ricardo Jorge. Given that data was irreversibly anonymised, the need for the participants’ informed consent was waived by the Ethical Committee.

### Italy

This study, based on routinely collected data, will not be submitted for approval to an ethical committee because the dissemination of COVID-19 surveillance data was authorized by the Italian law N. 52 of 19 May 2022, following the law decree N. 24 of 24 March 2022 (Article n. 13). Based on the same acts, the information on COVID-19 vaccination was retrieved by the Italian National Institute of Health using data from the National Immunisation Information System of the Italian Ministry of Health. Because of the retrospective design and the large size of the population under study, in accordance with the Authorization n. 9 released by the Italian data protection authority on 15 December 2016, the individual informed consent was not requested for the conduction of this study.

### Sweden

The Swedish study is approved by the Swedish Ethical Review Authority (2020-06859, 2021-02186) and has conformed to the principles embodied in the Declaration of Helsinki. Consent to participate is not applicable as this is a register-based study.

## Data Availability

Authors cannot share the data used for this study, which should be requested to the data owner institutions following their respective procedures.

## Author contribution

SB, NN, BN, JH and SM conceived the study and BN, JH, NN and SM conceited the methods. All authors from Public Health institutions at each study site were responsible for the data management and analysis at the site level. JH was in charge of pooling site estimates, with the help of SM. BN drafted the first version of the manuscript, with the help of JH and SM, AN, and EK. All the authors contributed to the interpretation of the results and critically reviewed the manuscript. All the authors approved the final version of this manuscript. All the authors within the VEBIS-EHR working group made a substantial contribution to the conception or design of the work, critically revised the manuscript, provided their final approval of the version to be published, and agreed to be accountable for all aspects of the work.

## References

1. European Centre for Disease Prevention and Control. Interim COVID-19 vaccination coverage in the EU/EEA during the 2023–24 season campaigns [Internet]. ECDC; 2024 [cited 2024 May 28]. Available from: https://www.ecdc.europa.eu/en/publications-data/interim-covid-19-vaccination-coverage-eueea-during-2023-24-season-campaigns

2. European Centre for Disease Prevention and Control. Communicable disease threats report, 17-23 December 2023, week 51. ECDC; 2023.

3. Hansen CH, Moustsen-Helms IR, Rasmussen M, Søborg B, Ullum H, Valentiner-Branth P. Short-term effectiveness of the XBB.1.5 updated COVID-19 vaccine against hospitalisation in Denmark: a national cohort study. Lancet Infect Dis. 2024 Feb 1;24(2):e73–4.

4. Monge S, Humphreys J, Nicolay N, Braeye T, Van Evercooren I, Holm Hansen C, et al. Effectiveness of XBB.1.5 Monovalent COVID-19 Vaccines During a Period of XBB.1.5 Dominance in EU/EEA Countries, October to November 2023: A VEBIS-EHR Network Study. Influenza Other Respir Viruses. 2024 Apr;18(4):e13292.

5. Caffrey AR, Appaneal HJ, Lopes VV, Puzniak L, Zasowski EJ, Jodar L, et al. Effectiveness of BNT162b2 XBB vaccine in the US Veterans Affairs Healthcare System. medRxiv. 2024 Jan 1;2024.04.05.24305063.

6. Tartof SY, Slezak JM, Puzniak L, Frankland TB, Ackerson BK, Jodar L, et al. Effectiveness of BNT162b2 XBB Vaccine against XBB and JN.1 Sub-lineages. medRxiv. 2024 Jan 1;2024.05.04.24306875.

7. Yang S, Yu Y, Xu Y, Jian F, Song W, Yisimayi A, et al. Fast evolution of SARS-CoV-2 BA.2.86 to JN.1 under heavy immune pressure. Lancet Infect Dis. 2024 Feb 1;24(2):e70–2.

8. Kirsebom FCM, Stowe J, Lopez Bernal J, Allen A, Andrews N. Effectiveness of autumn 2023 COVID-19 vaccination and residual protection of prior doses against hospitalisation in England, estimated using a test-negative case-control study. J Infect. 2024 May 7;89(1):106177.

9. Sentís A, Kislaya I, Nicolay N, Meijerink H, Starrfelt J, Martínez-Baz I, et al. Estimation of COVID-19 vaccine effectiveness against hospitalisation in individuals aged ≥ 65 years using electronic health registries; a pilot study in four EU/EEA countries, October 2021 to March 2022. Euro Surveill Bull Eur Sur Mal Transm Eur Commun Dis Bull. 2022 Jul;27(30).

10. Kislaya I, Sentís A, Starrfelt J, Nunes B, Martínez-Baz I, Nielsen KF, et al. Monitoring COVID-19 vaccine effectiveness against COVID-19 hospitalisation and death using electronic health registries in ≥65 years old population in six European countries, October 2021 to November 2022. Influenza Other Respir Viruses. 2023 Nov;17(11):e13195.

11. Fontán-Vela M, Kissling E, Nicolay N, Braeye T, Van Evercooren I, Holm Hansen C, et al. Relative vaccine effectiveness against COVID-19 hospitalisation in persons aged ≥ 65 years: results from a VEBIS network, Europe, October 2021 to July 2023. Vol. 29, Eurosurveillance. 2024. p. 2300670.

12. European Centre for Disease Prevention and Control. Protocol for a COVID-19 vaccine effectiveness estimation using health data registries, VEBIS multi-country study – Version 2.0 [Internet]. ECDC; 2024 [cited 2024 May 28]. Available from: https://www.ecdc.europa.eu/en/publications-data/protocol-covid-19-vaccine-effectiveness-estimation-using-health-data-registries

13. European Centre for Disease Prevention and Control. European Respiratory Virus Surveillance Summary (ERVISS) [Internet]. 2024 [cited 2024 Jun 27]. Available from: https://erviss.org/

14. Higgins JPT, Thompson SG. Quantifying heterogeneity in a meta-analysis. Stat Med. 2002 Jun 15;21(11):1539–58.

15. Moustsen-Helms IR, Bager P, Larsen TG, Møller FT, Vestergaard LS, Rasmussen M, et al. Relative vaccine protection, disease severity, and symptoms associated with the SARS-CoV-2 omicron subvariant BA.2.86 and descendant JN.1 in Denmark: a nationwide observational study. Lancet Infect Dis [Internet]. [cited 2024 Jun 20]; Available from: 10.1016/S1473-3099(24)00220-2

16. Hansen CH. Bias in vaccine effectiveness studies of clinically severe outcomes that are measured with low specificity: the example of COVID-19-related hospitalisation. Euro Surveill Bull Eur Sur Mal Transm Eur Commun Dis Bull. 2024 Feb;29(7).

