## Supplemental material for "Monovalent XBB.1.5 COVID-19 vaccine effectiveness against hospitalisations and deaths during the Omicron BA.2.86/JN.1 period among older adults in seven European countries: A VEBIS-EHR Network Study"

**Annex 6. Number of individuals, person-months and events, and VE estimate at study level .............14**

#### Annex 1. Methods to identify the BA.2.86/JN.1 predominance period, and study period:

To ensure robust identification of the BA.2.86/JN.1 predominance period, a minimum sample size threshold for the number of weekly specimens per country was determined based on the chosen predominance threshold (80%) and on limiting the coefficient of variation (CV) of the sub lineage proportion estimator to be 0.2. The sample threshold was calculated as: $n = (1-\hat{p})/(CV^2 \times \hat{p})$ where $\hat{p}$ is the predominance threshold and n is the minimum required sample size.

For countries with insufficient counts of sequenced cases, sub lineage proportions were imputed by pooling sequenced cases with those of neighbouring countries, defined by shared borders for contiguous nations and nearest 3 neighbours by centroid distance for islands.

Predominance periods were defined at the country level as starting in the first week in which the variant proportion exceeded the threshold (80% of all sequenced samples).

Figure S1. Weekly percentage of BA.2.86 samples in sequencing, per study site, and predominance period start date imputed (red) versus original ERVISS data (green).

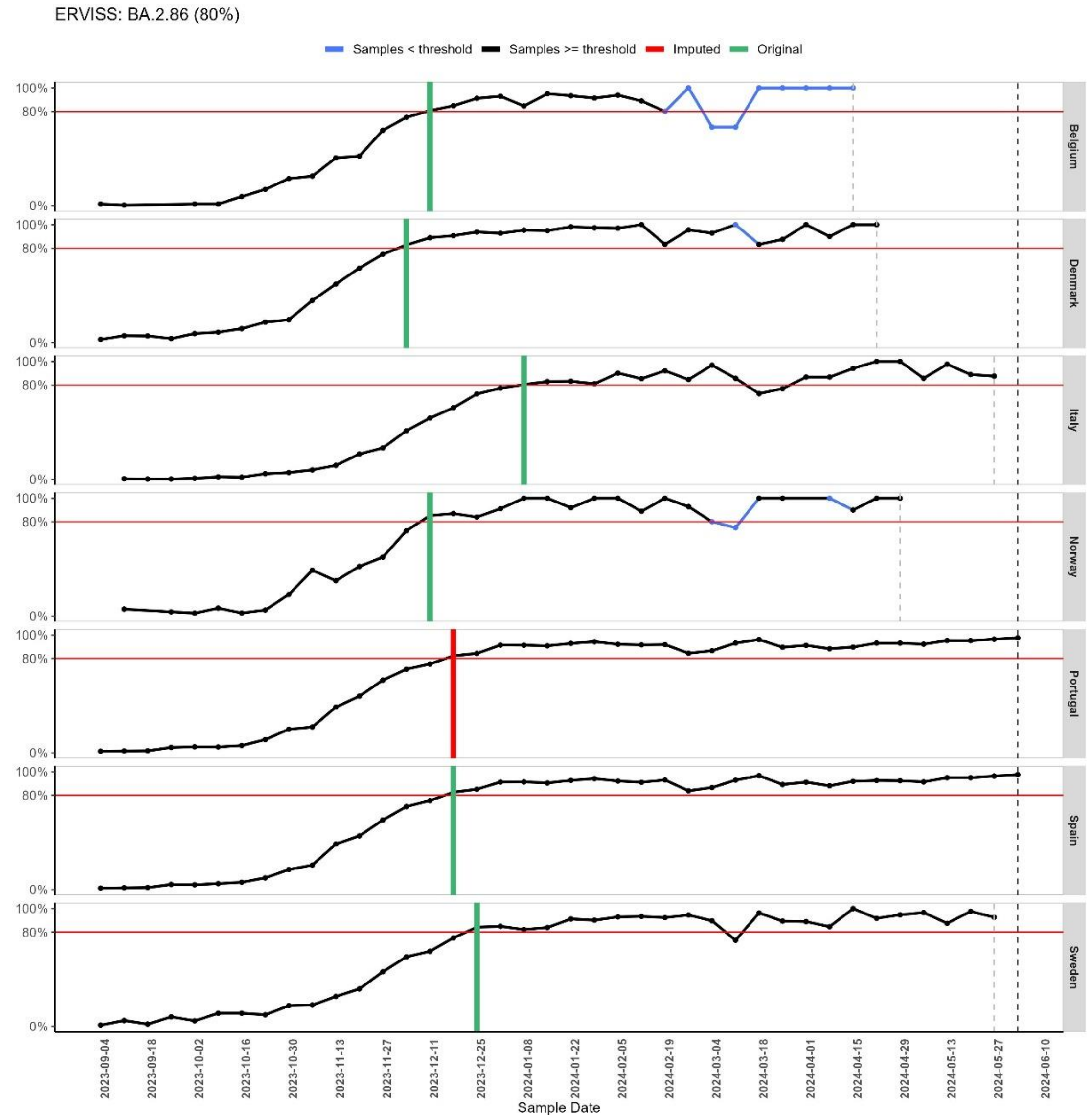

**Samples < Sample threshold are highlighted in blue, indicating that the number of sequenced BA.2.86 samples was below the required threshold (7 samples) for that week.*

Table S1. Start of the COVID-19 2023 autumnal vaccination campaign in participating countries

| **Study-site** | **Date of start of the 2023 autumn COVID-19 vaccination campaign** | **Start of study period (BA.2.86/JN.1 predominance period)*** ** |
|---|---|---|
| Belgium | 11 September 2023 | 11 December 2023 |
| Denmark | 01 October 2023 | 4 December 2023 |
| Italy | 27 September 2023* | 8 January 2024 |
| Navarre (Spain) | 25 September 2023** | 18 December 2023 |
| Norway | 1 September 2023 | 11 December 2023 |
| Portugal | 29 September 2023 | 18 December 2023 |
| Sweden | 1 September 2023 | 25 December 2023 |

* Day of publication of the Ministerial Circular Law with the recommendations on which vaccines to use and to whom they should be offered. The actual start day varied across regions, with most starting in the first week of October.

** The administration of the first autumnal doses began on 25th September. However, the campaign was officially launched on 16th October 2023 together with the flu campaign.

*** BA.2.86/JN.1 predominance period was defined as the set of weeks with BA.2.86/JN.1 variant weekly frequency equal or above 80% based on ERIVISS data.

#### Annex 2. Selection criteria according to the scientific protocol

The study population includes community-dwelling individuals ≥65 years of age in national and regional databases. The study population should be eligible for COVID-19 vaccination during the 2023 autumnal vaccination campaign, including belonging to an age group for whom autumnal COVID-19 vaccination has been recommended in each site/country. Eligibility will be based on the following criteria as of the first day of the vaccination campaign, which may be adapted to match national recommendations:

- Aged between 65 and 110 years at the beginning of the vaccination campaign, or belonging to an age group over 65 years for which the vaccine dose being evaluated has been recommended, if different (it should be recommended for the entire age group). Birth year may be used instead of age in countries where vaccine recommendations are based on birth cohort or only year of birth is available.
- Permanent resident in the EU/EEA territory covered in the study (for each study site, according to the most recent information when available).
- Not residents of a nursing home/long term care facility (according to the most recent information at the beginning of the autumnal vaccination campaign when available).
- Received their first ever COVID-19 vaccine dose as part of an age-specific vaccination campaign (i.e. excluding those vaccinated before it was generally recommended in the corresponding age group or, alternatively and at the discretion of each study site, excluding the first 5% of persons vaccinated within each age group – for each 5-year age bracket- as these first vaccinees may not be representative of their corresponding age group).
- Completed primary vaccination at least 180 days before the start of the autumnal vaccination campaign.

- Has not received a COVID-19 vaccine dose, irrespective of the number of doses, in the last 90 days before the start of the autumnal vaccination campaign; has no documented SARS-CoV-2 infection (nor has been hospitalised due to COVID-19) in the 90 days before the start of the autumnal vaccination campaign, or; other criteria following the relevant national guidelines of each study site (for example, if autumnal vaccine is recommended at ≥180 days after the last dose).
- Does not have inconsistent or missing data on vaccination (vaccination status unknown, any vaccination date is unknown, any vaccine brand is unknown, number of doses is unknown, interval between primary course first and second dose is shorter than 19 days, interval between complete primary vaccination and booster dose or between booster doses is shorter than 90 days, number of doses higher than recommended, received any vaccine brand not approved by EMA, or the combination of vaccine brands is not a recommended schedule -may vary by age group).

### Annex 3. Methodological details in the seven study sites

**Table S2. Data sources used in the six study sites to extract the study variables**

| Type of variables | Study variable | Study site | | | | | | |
|---|---|---|---|---|---|---|---|---|
| | | **Belgium** | **Denmark** | **Italy** | **Navarre (Spain)** | **Norway*** | **Portugal** | **Sweden** |
| **Outcomes** | **Hospital admission due to COVID-19** | Clinical Hospital Survey database | Danish National Patient Register (DNPR) | National Integrated COVID-19 Surveillance Databases | Enhanced COVID surveillance with individual revision of events | Norwegian Patient Register (NPR) | National Hospital Discharge database (BIMH) | Swedish National Patient Register |
| | **Death due to COVID-19** | Not applicable | MiBA and Danish Civil Registration system (CPR) | National Integrated COVID-19 Surveillance Databases | Administrative database of deaths and individual revision of events | Norwegian Death Registry (DÅR) | National Death Registry (SICO) and National Health Service User databaset (NHSU).Cause of death is from SICO, death status and date of death from NHSU. | Swedish Cause of Death register and Register of the Total Population Register on surveillance of notifiable communicable diseases |
| **Exposures** | **Vaccination status** | National vaccine registry (VACCINNET) | Danish Vaccination Registry (DVR) | National Vaccination Registry | Vaccination register | The National Immunisation Register (SYSVAK) | The National Vaccination Register (VACINAS) | Swedish National Vaccination Register |
| **Variables for adjustment or stratification** | **Age** | The national population register | CPR | National Vaccination Registry | Administrative database | The National Population Register (Folkeregisteret) | National Health Service User database (NHSU) | Register of the Total Population |
| | **Sex** | national population register | CPR | National Vaccination Registry | Administrative database | The National Population Register (Folkeregisteret) | National Health Service User database (NHSU) | Register of the Total Population |
| | **Health Region** | Province of residence: national population register | CPR | Region where vaccination took place National Vaccination Registry | Not applicable | County of residence at end of study period: The National Population Register (Folkeregisteret) | Region of residence: National Health Service User database (NHSU) | Register of the Total Population |
| | **Comorbidities** | Intermutualistic Agency database | DNPR | National Vaccination Registry | Primary Care clinical record | Risk groups / Comorbidities: Based on Norwegian Patient Registry (NPR) | Primary Care Information System (SIM@SNS). | Swedish National Patient Register |

| | | | | | | | | |
|---|---|---|---|---|---|---|---|---|
| | **Previous booster doses** | National vaccine registry (VACCINNET) | Danish Vaccination Registry (DVR) | National Vaccination Registry | Vaccination register | The National Immunisation Register (SYSVAK) | The National Vaccination Register (VACINAS) | Swedish National Vaccination Register |
| | **Others specific to the study site** | Household income (according to tax records) categorized as low (lowest 40%), mid (middle 30%), and high (highest 30%): STATBEL database | Not applicable | Country of birth National Vaccination Registry | Country of birth and high functional dependence: Administrative database | 1. Conditions of living – Crowding: Statistics Norway (SSB). Most recent data from 2019 – separate level for missing data<br>2. *County of birth:* Folkeregisteret | Conditions of living – Deprivation at the municipality level: Most recent data from Census 2011 | Marital status, educational level, country of birth The Longitudinal integrated database for health insurance and labour market studies |

*****All data in Norway was integrated in the emergency preparedness register for COVID-19 (Beredt C19), https://www.fhi.no/en/id/infectious-diseases/coronavirus/emergency-preparedness-register-for-covid-19/

**Table S3. Definition of co-variates, categorisation and use in the model**

| Variable | Definition, categorisation, and use in the model | | | | | | |
|---|---|---|---|---|---|---|---|
| | **Belgium** | **Denmark** | **Italy** | **Navarre (Spain)** | **Norway** | **Portugal** | **Sweden** |
| **Age** | Age in years at the end of the year in which the study period begins.<br>For adjustment: 5-year age groups. | adjusted in categories: 5-year categories until the final category, 90+ years | Age at the start of study period (for adjustment: 5-year age groups up to 90-94 years and then grouping ≥95 years)) | Age at the start of study period (5-year categories) | Age at end of 2023 (birth cohorts) (For adjustment: 5-year age groups) | Age at the start of study period (5-year categories) | Age in years at the end of the year in which the study period begins. |
| **Comorbidities** | No comorbidities associated with an increased risk for severe COVID-19 infection.<br><br>At least one comorbidity which increases the risk for severe COVID-19 infection and not being immunocompromised (medium risk):<br>- *Cardiovascular illness – general*<br>- *Cardiovascular illness-specifically a heart disease*<br>- *Alzheimer*<br>- *Asthma*<br>- *Haemophilia*<br>- *Chronic obstructive pulmonary disease*<br>- *Diabetes with cardiovascular compilations*<br>- *Diabetes Mellitus with insulin treatment*<br>- *Epilepsy and neuropathic pain*<br>- *Chronic hepatitis type B or C*<br>- *Exocrine pancreatic disease*<br>- *Disease of Parkinson*<br>- *Psychosis occurring with people older than 70 years*<br>- *Psychosis occurring with people of 70 year or younger.*<br>- *Thrombosis while treated with antithrombotic medicines*<br>- *Thyroid disorder*<br>- *HIV* | Immunocompromised, including:<br>- *HIV*<br>- *Immunological disease*<br>- *Radiation therapy*<br>- *Organtransplanted*<br><br>Other, including:<br>- *Diabetes*<br>- *Obesity*<br>- *Cancer*<br>- *Neurological Disease*<br>- *Kidney disease*<br>- *Haematological cancers*<br>- *Heart disease*<br>- *Chronic respiratory disease*<br>- *Liver disease (incl. alcohol lever)*<br>- *Endocrine Disease*<br>- *Hematological Disease*<br>- *Coagulation Disease*<br>- *Innate Diseases*<br>- *TB*<br>- *Missing a lung*<br>- *Missing a kidney* | Immunocompromised, including:<br>- *Immunocompromised defects of the complement system*<br>- *Other specified disorders involving the immune mechanism*<br>- *Deficiency or dysfunction of a single component (C1-C9)*<br>- *Deficiency of cell-mediated immunity*<br>- *Deficiency of humoral immunity*<br>- *Human immunodeficiency virus [HIV] disease, Human immunodeficiency virus, type 2 [HIV-2], Asymptomatic human immunodeficiency virus [HIV] infection status*<br>- *Disorders involving the immune mechanism*<br>- *Congenital and acquired disorders with poor antibody production*<br>- *Drug-induced immunosuppression*<br><br>Other comorbidities, including:<br>- *Respiratory diseases requiring oxygen therapy, idiopathic pulmonary fibrosis*<br>- *Advanced heart failure (Classes III-IV NYHA) and post cardiogenic shock patients*<br>- *Amyotrophic lateral sclerosis and other motor neuron disorders, multiple sclerosis, muscular dystrophy, infantile cerebral palsy, myasthenia* | Immunocompromised<br><br>Other major chronic conditions<br>- *Diabetes*<br>- *Severe Obesity*<br>- *Cancer*<br>- *Ictus*<br>- *Dementia*<br>- *Kidney disease*<br>- *Haematological cancers*<br>- *Heart disease*<br>- *Chronic respiratory disease*<br>- *Liver disease*<br>- *Rheumatic arthritis* | High risk:<br>- *Organ transplant*<br>- *Immunodeficiency*<br>- *Haematological cancer in the last five years*<br>- *Other active cancers*<br>- *Neurological or neuromuscular diseases that cause impaired cough or lung function (e.g., ALS and cerebral palsy)*<br>- *Chronic kidney disease, or significant renal impairment.*<br><br>Medium risk:<br>- *Chronic liver disease or significant hepatic impairment*<br>- *Immunosuppressive therapy*<br>- *Diabetes*<br>- *Chronic lung disease including cystic fibrosis and severe asthma which have required the use of high dose inhaled or oral steroids within the past year* | *Considered comorbidities include: anaemia, asthma, cancer, cardiac disease, dementia, diabetes, hypertension, HIV, liver disease, neuromuscular disease, obesity, pulmonary disease, renal disease, rheumatologic disease, stroke, tuberculosis* | *Vaccine priority groups:*<br>*LISA Healthcare worker (status per October 2018)*<br>*SOL Nursing home resident (status per December 31, 2020).*<br>*Comorbidity groups:*<br>*(binary)*<br>*Any records with ICD-10 codes as primary/secondary diagnosis from inpatient stay or outpatient contact in hospital or from privatepracticing specialists, January 1, 2017 – December 27, 2020)*<br>*Chronic pulmonary disease NPR J41 J42 J43 J44 J45 J46 J47 J84 J98 E84*<br>*Cardiovascular conditions and diabetes NPR, SPDR I05 I06 I07 I08 I09* |

| | | | | | | | |
|---|---|---|---|---|---|---|---|
| | Immunocompromised (high risk): if a person has one of the following comorbidities associated with immunodeficiency:<br>- *Disease of Crohn, Colitis Ulcerosa, Psoriatrische arthritis, Reumatoid arthritis*<br>- *Kidney failure*<br>- *Cystic fibrosis*<br>- *Psoriasis*<br>- *Multiple sclerosis*<br>- *Organ transplantation*<br>- *Received chemotherapy/radiotherapy against cancer*<br>- *Received multidisciplinary oncologic consult* | | *gravis, dysimmune neuropathies*<br>- *Type 1 diabetes, Type 2 diabetes with complications or requiring combination therapy (with at least two anti-diabetes drugs)*<br>- *Addison's disease*<br>- *Panhypopituitarism*<br>- *Cystic fibrosis*<br>- *-Cirrhosis of the liver*<br>- *Intracerebral ischemic or hemorrhagic event that has led to impaired neurological and cognitive autonomy*<br>- *Individuals who have had a stroke on 2020 or later ranked as level 3 or higher*<br>- *Thalassemia major*<br>- *Sickle cell anemia*<br>- *Other severe anemias*<br>- *Down syndrome*<br>- *Body Mass Index >35*<br>- *Severely disabled persons pursuant to law 104/1992 art. 3 paragraph 3*<br>- *Chronic Alcohol Misuse*<br>- *Functional or anatomic asplenia*<br>- *COPD*<br>- *Chemotherapy or Radiotherapy*<br>- *Coagulopathies*<br>- *Diabetes Mellitus and other endocrinopathies*<br>- *Patients in hemodialysis or with chronic kidney diseases expected to start dialysis*<br>- *Hemoglobinopathy such as sickle cell anemia or thalassemia*<br>- *Chronic Liver Disease*<br>- *Cochlear implant*<br>- *Chronic Kidney Disease*<br>- *Chronic eczema or psoriasis*<br>- *Diseases associated with a high risk of aspiration pneumonia*<br>- *Chronic Cardiovascular Disease*<br>- *Chronic Respiratory Disease*<br>- *Motor neuron diseases* | | - *Obesity with a body mass index (BMI) of ≥35 kg/m2*<br>- *Dementia*<br>- *Chronic heart and vascular disease (with the exception of high blood pressure) and stroke* | | *I110 I2 I34 I35 I36 I37 I39 I42 I43*<br>*I46 I48 I49 I50 E10-E14*<br>*ATC: A10 (at least two filled prescriptions during 2020, before December 27, 2020)*<br>*Autoimmunity-related conditions*<br>*NPR D86 G35 K50 K51 L40 M05 M06 M07 M08 M09 M13*<br>*M14 M45*<br>*Malignancy NPR, CAN C0 C1 C2 C3 C4 C5 C6 C7 C8 C9 D45 D46 D47 (CAN from 2017-2019, NPR for 2020)*<br>*Moderate to severe renal disease*<br>*NPR I12 I13 N00 N01 N02 N03 N04 N05 N07 N11 N14 N17 N18 N19 Q61* |

| | | | | | | | |
|---|---|---|---|---|---|---|---|
| | | | - *Chronic inflammatory diseases and malabsorption syndromes*<br>- *Blood cancers (leukemia, lymphoma and myeloma)*<br>- *Solid tumors*<br>- *Obesity (Body Mass Index 30-35)*<br>- *Bone marrow transplant*<br>- *Drug Misuse*<br>- *Solid organ transplant*<br>- *Patients with CSF leak from trauma or intervention*<br>- *Patients going to start immunosuppressive treatment*<br>- *Metabolic diseases*<br>- *Hematopoietic diseases*<br>- *Pathologies that require important surgical interventions*<br>- *Neurological diseases*<br>- *Cerebrovascular diseases*<br>- *Down Syndrome*<br>- *Disabilities (physical, sensorial, learning or psychic)* | | | | |
| **Country of residence / country of birth / nationality** | Not included | Not included | Country of birth: born in Italy; born in other countries | Country of birth | As registered at time of analysis (April 2024) | Not included | Country of birth. Sweden, Nordic, Eu, Other |
| **Deprivation index or similar** | Household income: low (lowest 40%)-medium (middle 30%)-high (highest 30%) | Not included | Not included | High functional dependence | Crowded conditions: if the number of rooms is lower than the number of residents or one resident lives in one room, and the number of square metres (P-area) is below 25 sq. m. per person. If the number of rooms or the P-area is not specified, a household was regarded as crowded if one of these criteria is met (incomplete and slightly outdated data) | European deprivation index quintile Q1 (least deprived) to Q5 (most deprived) | Individual education, marital status |

| | | | | | | | |
|---|---|---|---|---|---|---|---|
| **Geographic level** | Province of residence | Adjustment for residency in the 5 geographical regions of Denmark (EU NUTS-2 regions) | 19 regions and 2 autonomous provinces of Italy where vaccination took place | Not included | County of residence | Health region of residence (North, Center, Lisbon and Tagus Valey, Alentejo, Algarve) | Categories of rural/urban/metropolitan areas |
| **Number of COVID-19 tests in 2020-2022** | Not included | Positive RT-PCR test for SARS-CoV-2 | Not included | Not included | Not included | Not included | Positive RT-PCR test for SARS-CoV-2 |

#### Annex 4. Description of the variables included in the full confounding adjusted model per study site

Table S4: Full list of confounders included at each study site.

| Confounder | Belgium | Denmark | Italy | Navarre | Norway | Portugal | Sweden |
|---|---|---|---|---|---|---|---|
| Age group | X | X | X | X | X | X | X |
| Comorbidities | X | X | X | X | X | X | X |
| Previous booster doses | X | X | X | X | X | X | X |
| Sex | X | X | X | X | X | X | X |
| Region | X | X | X |  | X | X | X |
| Socioeconomic status | X |  |  |  | X | X | X |
| Country of birth |  |  | X | X |  |  |  |
| Dependence |  |  |  | X |  |  |  |

#### Annex 5. Number of individuals, person-months and events, and VE estimate at study level

**VE of autumnal vaccination, 65-79 years**

| Group / Study site | N | person-months | events | Fully adjusted VE (95% CI) |
|---|---|---|---|---|
| **Vaccinated (≥14 days ago; overall)** | | | | |
| Sweden | 981,663 | 1,990,555 | 133 | 52.0 [ 34.6, 64.8] |
| Portugal | 837,443 | 1,841,576 | 25 | 46.0 [ 7.5, 68.5] |
| Norway | 393,545 | 973,472 | 378 | 48.0 [ 39.3, 55.5] |
| Navarra | 53,892 | 149,933 | 32 | 45.3 [ 4.4, 68.7] |
| Italy | 783,586 | 1,188,159 | 55 | 49.0 [ 31.3, 62.2] |
| Denmark | 665,404 | 1,830,048 | 269 | 55.6 [ 43.0, 65.4] |
| Belgium | 792,028 | 1,988,972 | 16 | 60.0 [ 19.5, 80.1] |
| Pooled VE (fixed effects) | | | | 50.2 [ 44.6, 55.2] |
| Pooled VE (random effects) | | | | 50.2 [ 44.6, 55.2] |
| *Heterogeneity: I2 = 0%, tau2 = 0, p = 0.94* | | | | |
| **Vaccinated (≥14 and ≤89 days ago)** | | | | |
| Sweden | 975,599 | 1,634,950 | 128 | 52.0 [ 34.1, 65.0] |
| Portugal | 837,443 | 1,063,117 | 21 | 49.0 [ 10.2, 71.1] |
| Norway | 392,879 | 644,026 | 334 | 48.0 [ 38.8, 55.8] |
| Navarra | 53,592 | 107,980 | 26 | 52.4 [ 14.2, 73.6] |
| Italy | 780,595 | 984,842 | 47 | 50.0 [ 31.3, 63.6] |
| Denmark | 665,404 | 1,212,647 | 249 | 56.2 [ 43.7, 65.9] |
| Belgium | 791,772 | 829,368 | 8 | 62.0 [ 8.4, 84.2] |
| Pooled VE (fixed effects) | | | | 50.9 [ 45.1, 56.1] |
| Pooled VE (random effects) | | | | 50.9 [ 45.1, 56.1] |
| *Heterogeneity: I2 = 0%, tau2 = 0, p = 0.95* | | | | |
| **Vaccinated (90-179 days ago)** | | | | |
| Sweden | 722,760 | 355,605 | 5 | 67.0 [ 7.1, 88.3] |
| Portugal | 715,400 | 778,458 | <5 | |
| Norway | 341,873 | 329,445 | 44 | 50.0 [ 28.8, 64.9] |
| Navarra | 49,200 | 41,953 | 6 | -30.1 [-300.1, 57.7] |
| Italy | 328,164 | 203,316 | 8 | 37.0 [ -32.6, 70.1] |
| Denmark | 593,643 | 618,530 | 20 | 45.3 [ -0.4, 70.2] |
| Belgium | 765,507 | 1,159,605 | 8 | 58.0 [ -4.4, 83.1] |
| Pooled VE (fixed effects) | | | | 47.3 [ 32.0, 59.1] |
| Pooled VE (random effects) | | | | 47.3 [ 32.0, 59.1] |
| *Heterogeneity: I2 = 0%, tau2 = 0, p = 0.57* | | | | |

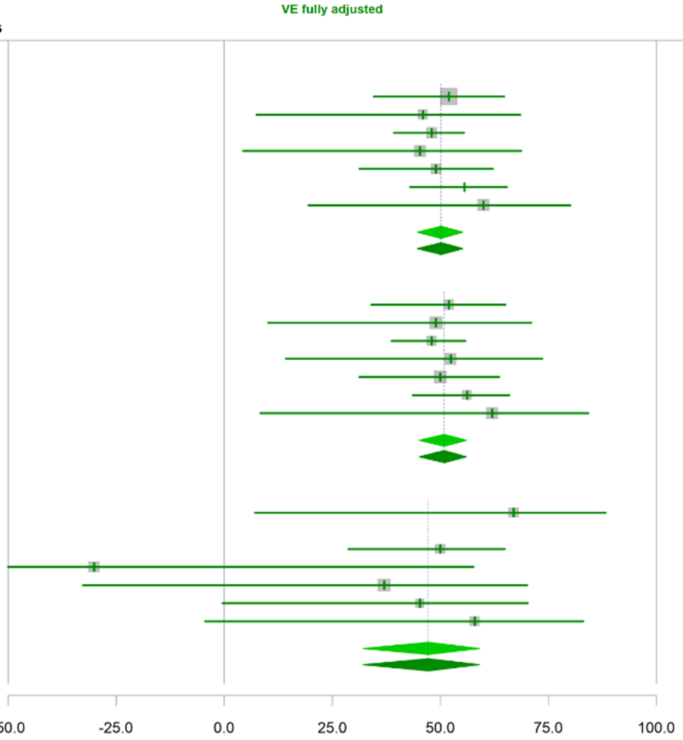

Figure S1 – Forest plots representing the study site (Belgium, Denmark, Italy, Navarre (Spain), Norway and Portugal) level number of persons, person-months, **COVID-19 hospitalisation events**, COVID-19 vaccine effectiveness estimates and respective 95% confidence interval, plus common COVID-19 VE estimate and respective 95% confidence interval, using the random and fixed effects model, including the measures of heterogeneity between vaccine hazard ratios between study sites $I^2$, $\tau^2$ and the p-value of Q Cocran test, for those with 14 or more days, 14-89 days and 90-179 days since vaccination, during the BA.2.86/JN.1 lineages predominance period, **among individuals aged between 65 to 79 years of age.** VEBIS-EHR network

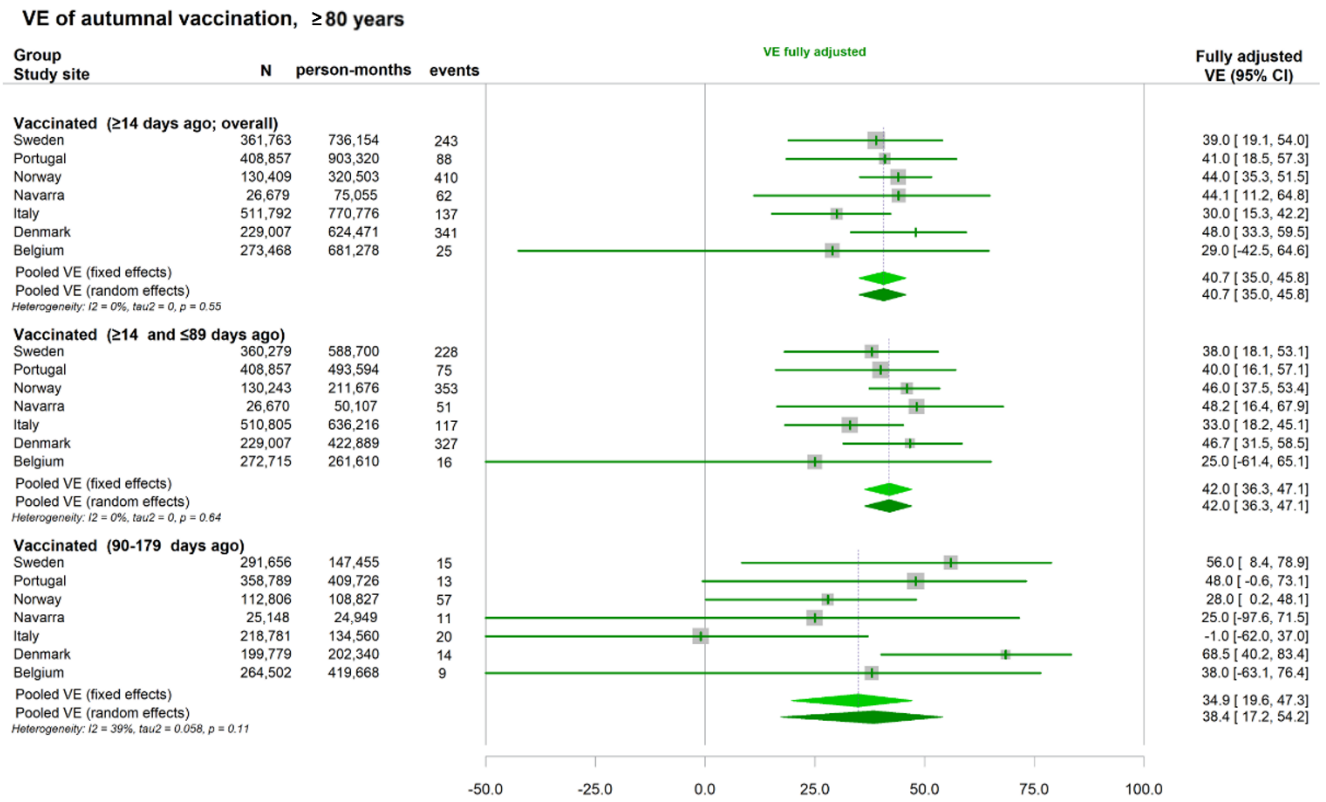

Figure S2 – Forest plots representing the study site (level number of persons, person-months, **COVID-19 hospitalisation events**, COVID-19 vaccine effectiveness estimates and respective 95% confidence interval, plus common COVID-19 VE estimate and respective 95% confidence interval, using the random and fixed effects model, including the measures of heterogeneity between vaccine hazard ratios between study sites $I^2$, $\tau^2$ and the p-value of Q Cocran test, for those with 14 or more days, 14-89 days and 90-179 days since vaccination, during the BA.2.86/JN.1 lineages predominance period, **among individuals aged ≥80 of age.** VEBIS-EHREHR network**.**

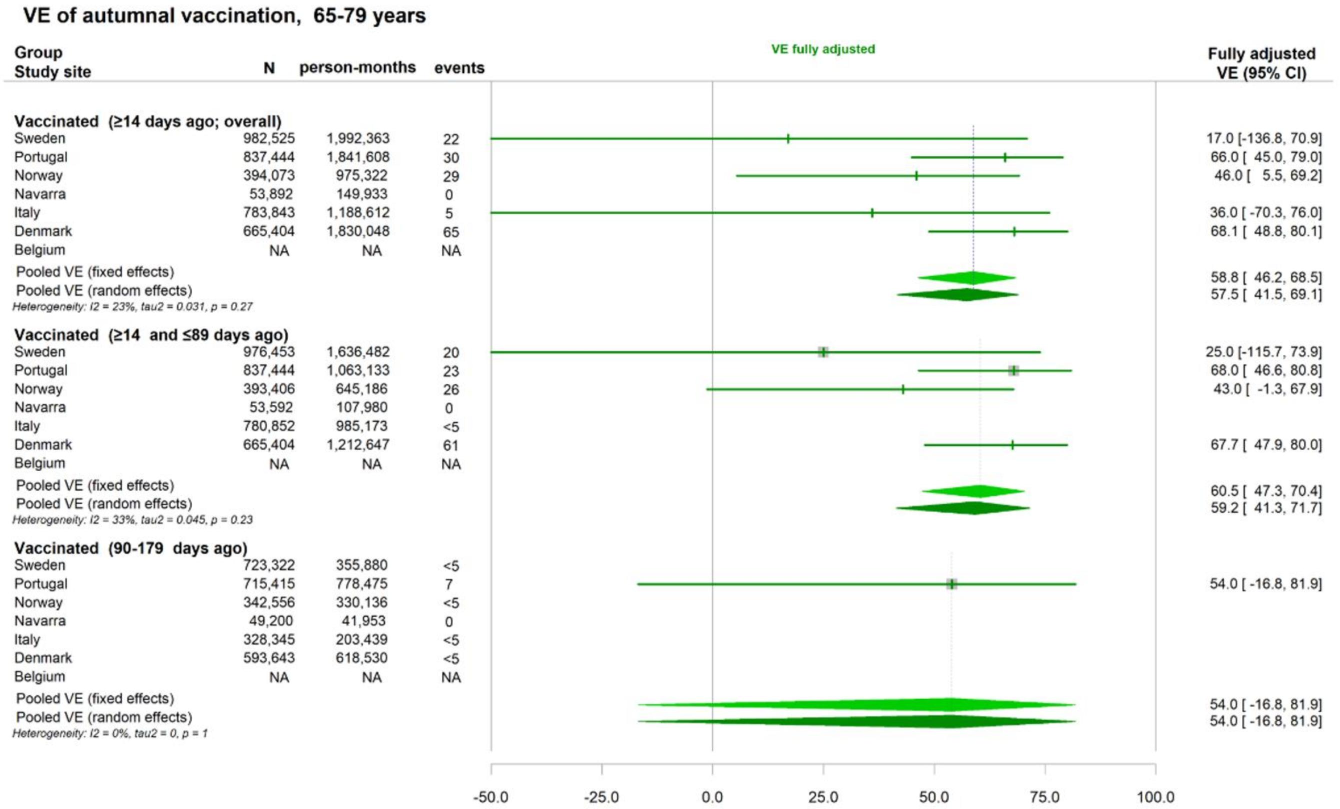

Figure S3 – Forest plots representing the study site level number of persons, person-months, **COVID-19 related death events**, COVID-19 vaccine effectiveness estimates and respective 95% confidence interval, plus common COVID-19 VE estimate and respective 95% confidence interval, using the random and fixed effects model, including the measures of heterogeneity between vaccine hazard ratios between study sites $I^2$, $\tau^2$ and the p-value of Q Cocran test, for those with 14 or more days, 14-89 days and 90-179 days since vaccination, during the BA.2.86/JN.1 lineages predominance period, **age among individuals aged between 65 to 79 years of age.** VEBIS-EHR network**.**

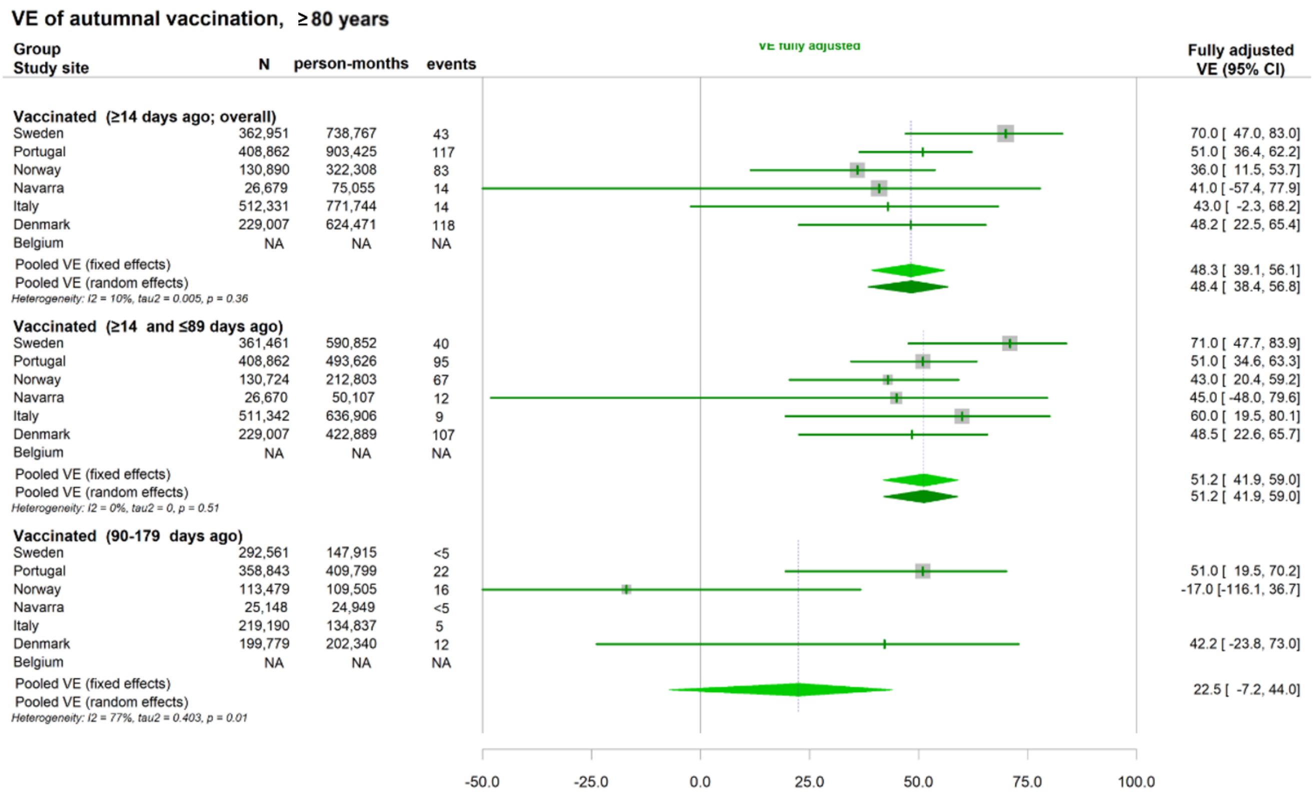

Figure S4 – Forest plots representing the study site level number of persons, person-months, **COVID-19 related death events**, COVID-19 vaccine effectiveness estimates and respective 95% confidence interval, plus common COVID-19 VE estimate and respective 95% confidence interval, using the random and fixed effects model, including the measures of heterogeneity between vaccine hazard ratios between study sites $I^2$, $\tau^2$ and the p-value of Q Cocran test, for those with 14 or more days, 14-89 days and 90-179 days since vaccination, during the BA.2.86/JN.1 lineages predominance period, **among individuals aged ≥80 of age.** VEBIS-EHR network.

#### Annex 6. COVID-19 2023 autumnal vaccine coverage within the study participants:

Table S5 – Estimates of COVID-19 Vaccine coverage in the VEBIS-EHR cohorts compared with estimates provided by countries in ECDC report (April 2024).

| | Vaccine coverage VEBIS-EHR n/N (%) | | Vaccine coverage (ECDC) | | |
|---|---|---|---|---|---|
| Age group | 65-79 | ≥80 | 60-69 | 70-79 | ≥80 |
| Study site | | | | | |
| Belgium | 792,060/1,544,854 (51.3) | 273,501/457,290 (59.8) | 37.3 | 55.9 | 57.6 |
| Denmark | 665,404/837,994 (79.4) | 229,007/275,898 (83.0) | 43.8 | 80.7 | 86.6 |
| Italy | 783,920/8,190,663 (9.6) | 512,544/3,856,362 (13.3) | 6.0 | 11.6 | 8.8 |
| Navarre (Spain)* | 102,792/138,810 (74.1) | 51,818/61,363 (84.4) | 32.6 | 52.3 | 64.3 |
| Norway | 394,126/699,402 (56.4) | 130,922/222,886 (58.7) | 30.3 | 61.2 | 61.8 |
| Portugal | 838,447/1,632,452 (51.4) | 412,644/719,488 (57.4) | 43.5 | 59.6 | 63.9 |
| Sweden | 983,389/1,365,267 (72.0) | 363,150/438,919 (82.7) | 46.8 | 73.8 | 89.3 |

* ECDC report estimates correspond to Spain and not Navarre only.
